# “Intellect”, a Mobile Health Application based on cognitive-behavioral therapy principles, improves Anxiety and Worry: A Randomized Controlled Trial with an Active Control and a 2-Week Follow-Up

**DOI:** 10.1101/2022.07.25.22278034

**Authors:** Feodora Roxanne Kosasih, Vanessa Tan Sing Yee, Sean Han Yang Toh, Oliver Suendermann

**Author notes:** Corresponding author (OS). These authors contributed equally to this work (Feodora Roxanne Kosasih, Vanessa Tan Sing Yee).

## Abstract

Digital self-guided mobile health (mHealth) applications are cost-effective, accessible, and well-suited to improve mental health at scale. This randomized controlled trial (RCT) evaluated the effectiveness of a recently developed mHealth programme based on cognitive-behavioral therapy (CBT) principles in improving worry. We also examined psychological mindedness (PM) as a mediator by which app engagement is thought to improve outcomes. The intervention group completed a 2-week “Anxiety and Worry” programme with daily CBT-informed activities, while the active waitlist-control completed a matched 2-week mHealth programme on procrastination. Participants filled out the Generalized Anxiety Disorder (GAD-7), Patient Health Questionnaire (PHQ-9), and Psychological Mindedness Scale (PMS) at baseline, post-intervention and 2-week follow-up. App engagement was measured at post-intervention only. Both groups showed significant improvements on anxiety and depression scores from baseline to post-intervention, but no group differences were observed. From post-intervention to follow-up, only the intervention group showed further improvements for anxiety levels. Higher engagement with the mHealth app reported lower anxiety at post-intervention, and this relationship was fully mediated by psychological mindedness. This study provides evidence that (a) engaging in a CBT mHealth App can effectively reduce anxiety and worry, and (b) Psychological mindedness is a potential pathway by which engaging with a mHealthapp improves worry. While overall effect sizes were small, at the population level, these can make significant contributions to public mental health.

**Author Summary:** Increasing burden of anxiety amongst young adults has made widely accessible mobile health applications a promising tool in improving anxiety levels at scale. We conducted a randomized controlled trial (N=309) to examine the effectiveness of a brief, publicly available mobile health application (Intellect’s “Anxiety and Worry” programme) in reducing anxiety and worry levels among young adults. Participants who received the intervention showed significant reduction in anxiety and depression levels, however, effects did not significantly differ from active control. At post-intervention, only the intervention group continued to experience improvements in anxiety level. We also found that higher app engagement with the mHealth app predicted better anxiety and depression outcomes, and this relationship was fully mediated by psychological mindedness. Future work would benefit from inclusion of waitlist control, a larger sample size, and identification of alternative mediators.

## Introduction

Worry is commonly defined as a chain of thoughts and images about future events that are negatively affect-laden and experienced as relatively uncontrollable (1). Persistent and excessive worry, also known as anxiety, is associated with an increased risk for cardiovascular illness (2), mortality and suicide (3), as well as reduced quality of life (2). University students are particularly vulnerable to anxiety, with an accumulated prevalence rate of 41% during the pandemic (4). While effective psychological interventions exist for worry, only few are utilizing these due to multiple barriers, such as high treatment costs, limited accessibility, lack of perceived need for help, and negative stigma (5–8). Unfortunately, severe worry usually takes a chronic course, and is unlikely to resolve without appropriate interventions (9). Therefore, there is a dire need for an effective, cost-efficient intervention on anxiety.

One solution to help large numbers of people is to utilize widely-accessible, secure yet inexpensive mobile health (mHealth) applications (10–12). Emerging research has found evidence that engaging with mHealth apps can improve anxiety across both clinical (10) and non-clinical populations (13,14). To date, several meta-analyses have highlighted promising evidence on the effectiveness of mHealth apps in decreasing anxiety using randomized controlled trials (RCT) which found small but significant effects (g = 0.30-38) (11,15). However, comparisons against active control often yield significantly weaker and inconsistent results (11,15,16). Nevertheless, scaled at population level, small effects can meaningfully impact public mental health.

Despite promising evidence on the effectiveness of mHealth apps on improving anxiety, especially for mild symptom severity, there is still a limited number of applications with reported effectiveness (17,18), quality (19), and evidence-based components (20). Wang et al. (18)’s systematic review found that only 14 out of over 100 studies had reported clinically validated evidence on its effectiveness. In a review of 27 popular apps by Wasil et al. (20), most apps contained just 3 evidence-based components and incorporated limited core treatment techniques typically present in standard anxiety treatment protocols (i.e. cognitive structuring made up of just 12% of the application). Therefore, there is a strong need for more empirical studies to gain a better perspective of the true value of mHealth apps on anxiety, paying extra attention to the content used within the application and the type of control groups used in the study.

While more efforts have been made to examine the effectiveness of publicly available mHealth apps, limited studies have investigated the underlying mechanisms that enable such impact. To our knowledge, only 5 studies evaluated such mechanisms (13,21–24), and only 3 of these studies utilized mHealth apps incorporating multiple Cognitive Behavior Therapy (CBT) components. Using *MoodMission, MoodPrism, and MoodKit*, Bakker and colleagues (13,21,22) demonstrated the mediating effect of coping self-efficacy in the relationship between app engagement ratings and anxiety, depression, and mental wellbeing outcomes. Bakker and Rickard (21) also found a partially mediated effect of emotional self-awareness (ESA) in the aforementioned relationship. Identifying additional mechanisms that facilitated improvements in mental health outcomes can further inform mHealth apps developers to create apps that utilize multiple mediators to enhance mental health outcomes (21). One such potential mediator is psychological mindedness (PM), defined by Denollet and Nyklíček (25) as “the intrinsic motivation to be in touch with one’s inner feelings and thoughts by monitoring and analyzing them in an adaptive way.” An important factor in the development of emotion regulation, PM has previously been linked to lower psychological distress and better coping abilities (26–33). Previous research has also hinted at the prospect of PM as a relevant skill in psychotherapy, albeit inconsistent findings on its effect on treatment outcomes (28,34–36). As psychotherapies encourage individuals to monitor, evaluate, and express their cognitions, emotions, and behaviors, it is likely that participating in treatments increases an individual’s PM (37–39). A study by Boylan (37) found an improvement in PM over time across treatments, including CBT. Furthermore, Nyklíček et al. (38) demonstrated that changes in PM scores over the course of an intervention were associated with larger decreases in psychological symptoms scores, including anxiety, depression, and sleeping problems in a clinical sample. Additionally, previous research has indicated that metacognitive skills such as PM are systematically developed during the transition from adolescence to adulthood and are considered key mechanisms required to transform behaviors during cognitive-behavior-based and mindfulness-based treatments (40,41). Altogether, this suggests that PM might act as a mediator that intermediately affects treatment outcomes from the use of mHealth apps.

### Objectives

This study aimed to evaluate the effectiveness of a recently developed mHealth application (“Intellect”) “Anxiety and Worry,” compared to an active control condition (“Procrastination”) among young adults in a randomized controlled trial (RCT) with a 2-week follow-up. We hypothesized that participants in the intervention group would experience significant decreases in anxiety (primary outcome) and depression (secondary outcome) compared to the active control condition. Based on Bakker and colleagues’ (13,21,22) prior work, we also investigated whether PM mediates the hypothesized relationship between engagement with the mHealth app and improvements in anxiety and depression scores.

As such, the current study aims to (1) confirm existing research on the effectiveness of a mHealth app in improving anxiety and depression outcomes by conducting a RCT with an active-control group and 2-week follow-up, and (2) explore PM as a possible mediator in improving anxiety and depression outcomes.

## Materials and Methods

### Design

This randomized controlled trial (RCT) is a 2 × 3 mixed factorial experimental design, with condition (intervention vs. active control) as the between-groups factor and time of assessment (pre-intervention vs. post-intervention vs. 2-week follow-up) as the within-groups factor. The study was registered with ClinicalTrials.gov (NCT04911803) and it was approved by the National University of Singapore’s Institutional Review Board NUS-IRB (Reference Code: 2021-266). The study was conducted entirely remotely. English-speaking participants aged 18 years and above were recruited between June 2021 and December 2021 through the University’s internal Research Recruitment portal, and through the Department of Psychology’s Research Participation (RP) pool. Participants were awarded either $15 reimbursement or 3 course credits for their participation. Sample characteristics for the final sample of 309 participants (M_age_= 22.05; SD_age_= 4.06, range: 18 to 54 years) are displayed in Table 4.

### Materials

#### “Anxiety and Worry” Programme (Intervention Condition)

This is a 14-day mHealth program that provides psychoeducation on the negative core beliefs and/or thought patterns associated with anxiety and worry, as well as effective coping mechanisms and problem-solving techniques. Each psychoeducation module is followed by short writing exercises, and a few situational-task questions (see Table 1 for a program overview and Table 3 for techniques utilized). This module structure allows participants to actively practice utilizing the techniques they have learned.

**Table 1.** Overview of the Anxiety Programme.

| Topics | Content |
| --- | --- |
| Understanding your worries | • Understanding Anxiety & Worry<br>• Learning how to spot Anxiety and what triggers it<br>• Understanding Core Beliefs and how it was formed<br>• Understanding your Coping Mechanism |
| Identifying your worries | • Introduction of the Worry Tree Technique |
| Working through your anxiety | • Develop skills to focus on the possible solutions of one's Anxiety |
| Rewiring your thought patterns | • Understand what thought patterns are and how to change them<br>• Recap of the entire programme and key information in each session |

**Table 2.** Overview of the Procrastination Programme.

| Topics | Content |
| --- | --- |
| Motivations behind procrastination | • Understanding procrastination and its cycle<br>• Learning what mental barriers are, how to spot and how to challenge it |
| Overcoming the barrier | • Learning and practicing mindfulness<br>• Identify source of discomfort that leads to procrastination<br>• Learning different ways to tolerate discomfort |
| Creating better habits | • Develop skills to create healthier habits<br>• Recap of the entire programme and key information in each section |

**Table 3.** Overview of Evidence-Based Treatment Components Covered.

| Treatment Component | Example Sub-Components | Anxiety Programme | Procrastination Programme |
| --- | --- | --- | --- |
| Psychoeducation | Education on Topic (definition, cycle of reinforcement, etc) | ✓ | ✓ |
| Self-Monitoring | Monitoring Cognition, Emotions, and Behaviors | ✓ | ✓ |
| Cognitive Techniques | Identifying Thoughts, Cognitive Restructuring Exercises | ✓ | ✓ |
| Journaling | Reflective Writing, Informative Journaling | ✓ | ✓* |
| Behavioral Techniques | Behavioral Activation, Behavioral Experiments | ✓* | ✓ |
| Problem Solving | Problem Solving Exercise | ✓ |  |
| Coping Strategies | Identifying coping abilities | ✓ |  |
| Relaxation Skills | Mindfulness Exercise |  | ✓ |

\*indicates minimal presence in the programme

**Table 4.** Descriptives of Primary and Secondary Measure Means and SDs at Baseline.

| Variables |  | Intervention Condition | Active Control Condition | *p-value* |
| --- | --- | --- | --- | --- |
|  |  | *M (SD)* | *M (SD)* |  |
| Age |  | 22.117 (4.282) | 21.974 (3.848) | 0.758 |
| GAD-7 |  | 7.701 (5.088) | 7.252 (5.422) | 0.453 |
| PHQ-9 |  | 7.805 (5.724) | 7.787 (5.928) | 0.978 |
| PMS |  | 126.097 (10.742) | 126.768 (10.874) | 0.586 |
| AES** |  | 27.330 (3.787) | 27.248 (3.804) | 0.878 |
|  |  | *N (%)* | *N (%)* |  |
| Gender | Female | 122 (79.22%) | 109 (70.32%) | 0.191 |
|  | Male | 31 (20.13%) | 45 (29.03%) |  |
|  | Others | 1 (0.65%) | 1 (0.65%) |  |

GAD-7 = Generalized Anxiety Disorder; PHQ-9 = Patient-Health Questionnaire; PMS =
intervention

#### “Procrastination” Programme (Active Control Condition)

The 14-day mHealth program on Procrastination provides psychoeducation on the procrastination cycle, the mental barriers that arise due to procrastination, and ways to tolerate and overcome the discomfort. Participants engaged daily in either short situational-task questions, short writing activities, or mindfulness activity. The structure and duration of the Procrastination Programme (PP) is similar to that of the Anxiety and Worry programme (see Table 2 for program overview and Table 3 for technique utilized).

### Procedures

Participants signed up for the study through the University’s recruitment sites, where they were redirected to a secured Qualtrics survey link. Participants who provided digital consent to participate in the study (N = 492) were prompted to complete the baseline questionnaires, where outcome variables (PHQ-9, GAD-7, PM) were measured. Only participants who were able to download the application, has not participated in similar mHealth app studies, and completed the baseline measures (N = 323) were then randomized to the intervention (“Anxiety and Worry” Programme N = 160) or the active control (“Procrastination” Programme N = 163) condition. We used partial concealment of the main purposes of the study by informing participants that the purpose of the study was to evaluate the benefits of using a mHealth app to improve wellbeing. Thereby we aimed to minimize the pygmalion effect, which is the phenomenon in which one’s expectation of a desired behavior leads to an improvement in that particular behavior. Next, participants were guided to download the Intellect mHealth application and were provided a unique code to register and access a research version of the app, where only their assigned programme was available. This was done to ensure that any treatment effects observed were due to the programme, and not other features available in the application. Both programmes span over 14 days, with one session to be completed daily (i.e., <5 minutes). After completion of their allocated mHealth programmes, participants were contacted to complete the same outcome variables (PHQ-9, GAD-7, PM) and the app engagement scale (AES). The same measures (but without the AES) were administered 14 days after the completion, and participants were also debriefed about the main purposes of the study.

### Measures

#### Primary Outcome Measures

##### Generalized Anxiety Disorder (GAD-7)

Generalized Anxiety Disorder-7 (GAD-7) is a 7-item self-report instrument that measures anxiety as a continuum (42) and is widely used in research in both clinical and nonclinical populations (43). Items are scored on a 4-point scale, ranging from Not at All (0) to Nearly Every Day (3). The reliability of GAD-7 in the present study was excellent (*α* = .84 to *α* = .90).

#### Secondary Outcome Measures

##### Patient Health Questionnaire (PHQ-9)

Patient Health Questionnaire (PHQ-9) is a 9-item self-report instrument that measures depressive symptomatology. Items are scored on a 4-point scale, ranging from Not at all (0) to Nearly Every Day (3), where higher scores indicate more depressive symptoms. The reliability of PHQ-9 in this study was very good (*α* = .85 to *α* = .86).

#### Psychological Mindedness Scale (PM)

Psychological Mindedness Scale (PM) is a 45-item self-report instrument that measures an individual’s ability to be reflective about interpersonal relationships, psychological processes, and meanings across both intellectual and emotional dimensions. Items are scored on a 4 point-scale ranging from Strongly agree (4) to Strongly Disagree (1). Consistent with previous research where *α* = .86 to *α* = .87 (28), the reliability of PM in this study was acceptable (*α* = .77 to *α* = .79).

#### App Engagement Scale (AES)

App Engagement Scale (AES) is a 7-item questionnaire based on the Mobile Application Rating Scale’s (MARS) items and themes (46). All 7 items are scored on a 5-point likert scale ranging from Strong disagree (1) to Strong agree (5). The AES displayed good internal reliability (*α* = .83).

### Statistical Analysis

#### Power Analysis

Power was calculated with G*Power v.3.1 (47). Using a small effect size from similar mHealth RCT’s (13,48), setting Cronbach alpha at .05, and the power at .90, N=88 participants per group was required. Accounting for high dropout rates observed in previous studies (13,49), we aimed to recruit a minimum of N=200 participants.

#### Data Screening

Data screening included examining the data for incompleteness. Missing data from the post-intervention and follow-up assessment were replaced with data from the last completed questionnaire to ensure data was analyzed as intention-to-treat (ITT).

All data were analyzed using IBM SPSS v.25. Preliminary data analyses included normality testing and checking for univariate outliers. 2 outliers (i.e. +/- 3SD from the mean) were excluded. T-tests and chi-square tests examined baseline group differences.

#### Main Analysis

Repeated-Measures Analysis of Covariance (RM ANCOVA) with time as within subject factor and condition (Anxiety, Procrastination) as between subject factors assessed whether participants reported a decrease in anxiety and depression from baseline (T1) to post-intervention (T2) and from post-intervention (T2) to follow-up (T3). In all models, programme completion duration and numbers of other features engaged in the application outside of the assigned programmes (from T1 to T2) were included as covariates.

Next, a series of mediation analyses were performed using Hayes’ PROCESS macro plugin Model 4 with confidence intervals set at 95% using bootstrapping procedures, with 5000 samples (95% confidence interval) (50). Four mediated regression models were used to examine the relationship between App Engagement Ratings and outcome variables, including Anxiety (GAD-7) and Depression (PHQ-9). The mediating variable used in the model was PMS sores at the respective times.

The main analyses were performed on the whole ITT sample (n=309), with the respective outcome measure at pre-intervention as covariates. The mediation analyses were performed on the ITT sample who completed the AES scale (n=201). Eta squared partial (η^2^_partial_ ≈ .01: small; η^2^_partial_ ≈ .06: medium; η^2^_partial_ ≈ .14: large) was the effect size reported for all findings (51).

## Results

### Participants

The CONSORT Flow Diagram is shown in Fig 1. No significant baseline group differences on baseline outcome measures, gender, and age, were observed (p>.05). There were no significant differences in application engagement (p>.05). Table 4 shows the sample characteristics and baseline scores.

**Fig 1.**
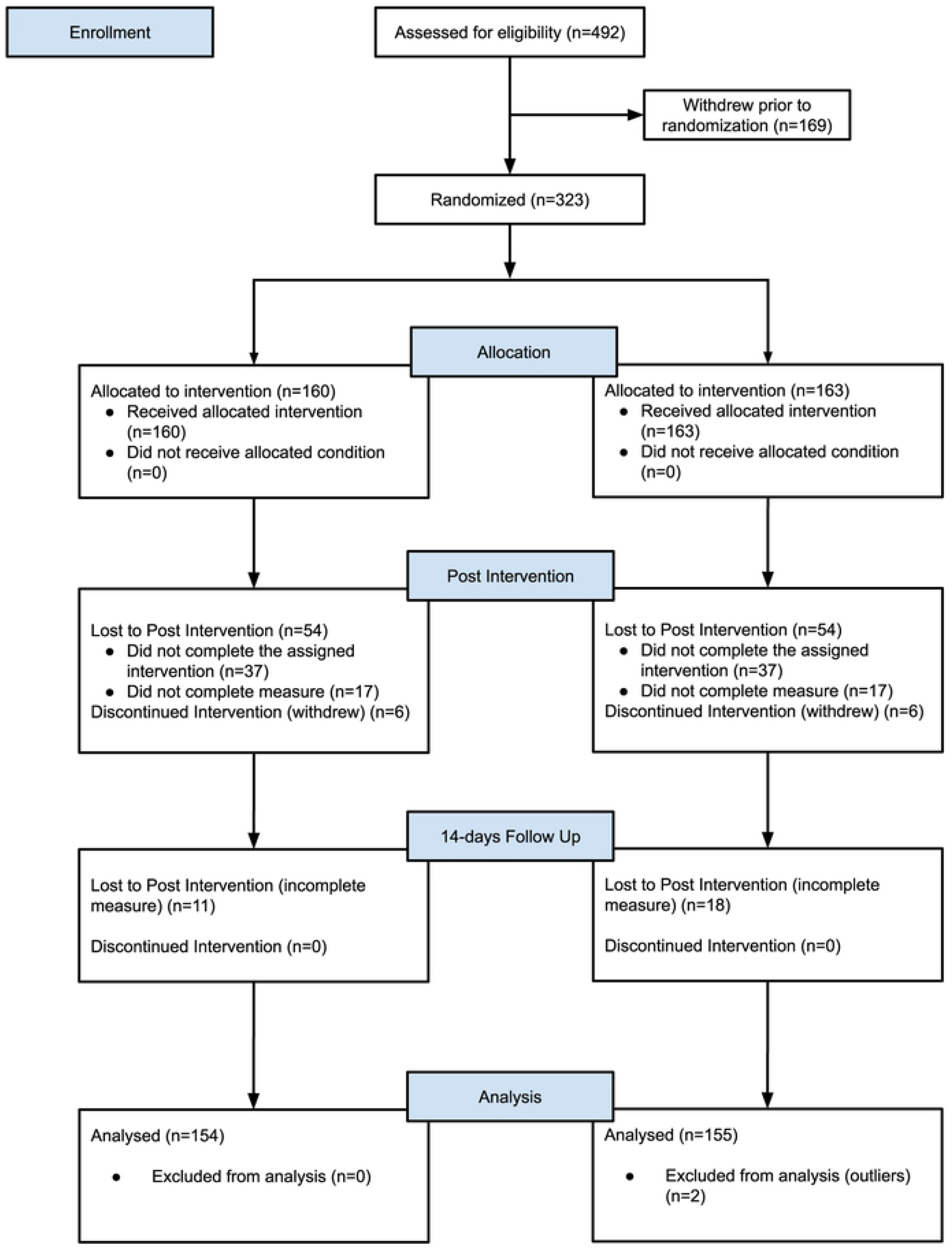
CONSORT Flow Diagram.

### Test of Group Effects

The repeated measures ANCOVA with anxiety scores as dependent variable revealed a significant main effect of time [F(1,307)=12.211, p<0.001, ηp^2^=0.038] and no significant interaction of time x condition [F(1,307)=2.055, p=0.153, ηp^2^=0.007], indicating a decrease in anxiety scores at post-intervention in both conditions. A similar significant main effect of time was also observed with depression scores as dependent variables, [F(1,307)=19.982, p<0.001, ηp2=0.061]. This indicates a decrease in depression scores at post-intervention in both conditions. No significant interaction of time x condition was observed [F(1,307)=0.625, p=0.430, ηp^2^=0.002].

From post-intervention (T2) to follow-up (T3), a significant interaction effect of time x condition [F(1,306)=4.855, p=0.028, ηp^2^=.016] was observed (see Fig 2), indicating the groups differed on how much their anxiety scores changed across both time points. Visual inspection indicated a trend for anxiety to decrease more strongly after completing the Intervention, while the Active Control condition did not experience such decline. The full results of the analyses are presented in Table 5.

**Table 5.**
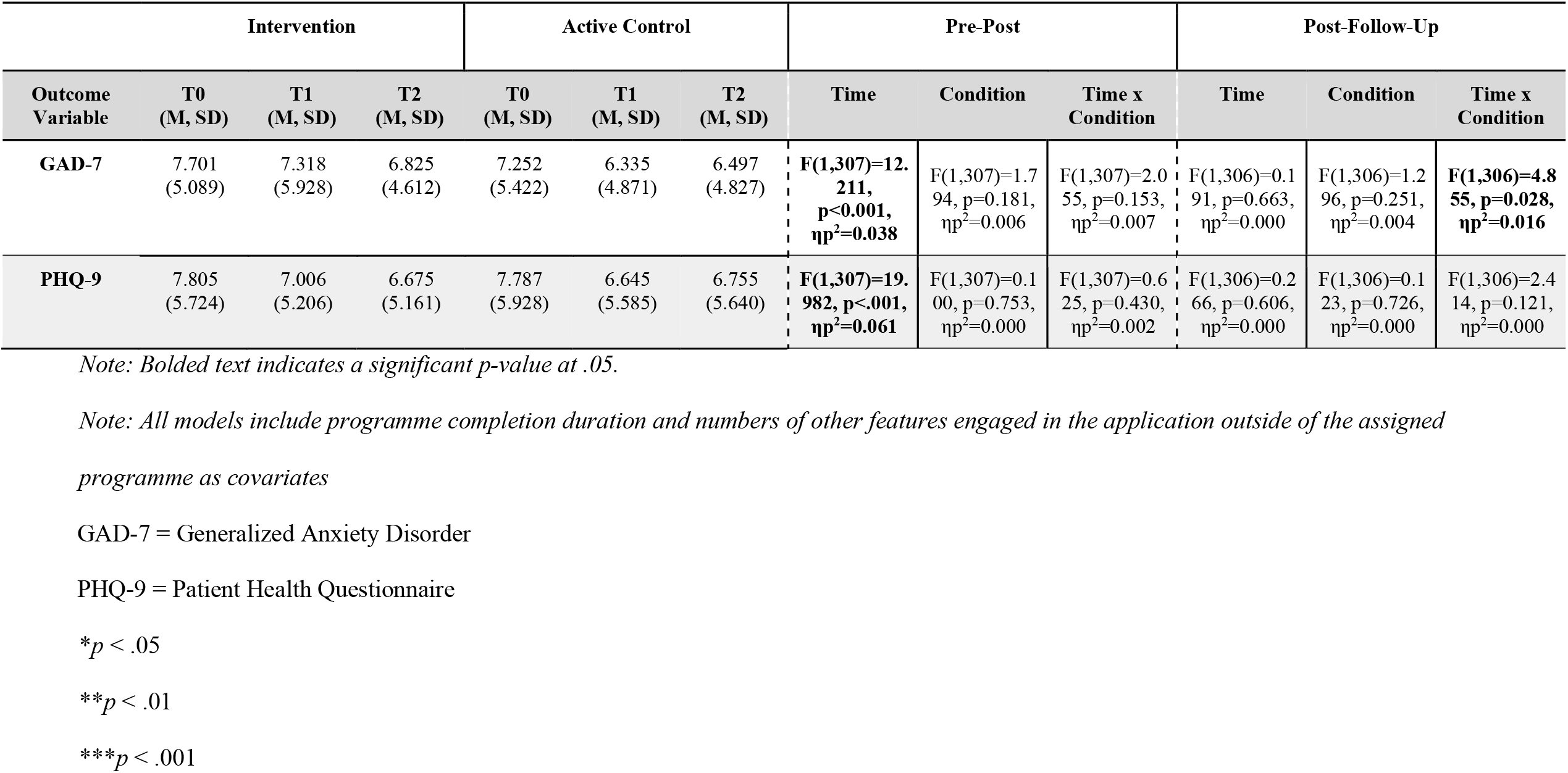
Comparison of Group Effects for Anxiety and Depression at post-intervention (T1-T2) and follow-up (T2-T3)

**Fig 2.**
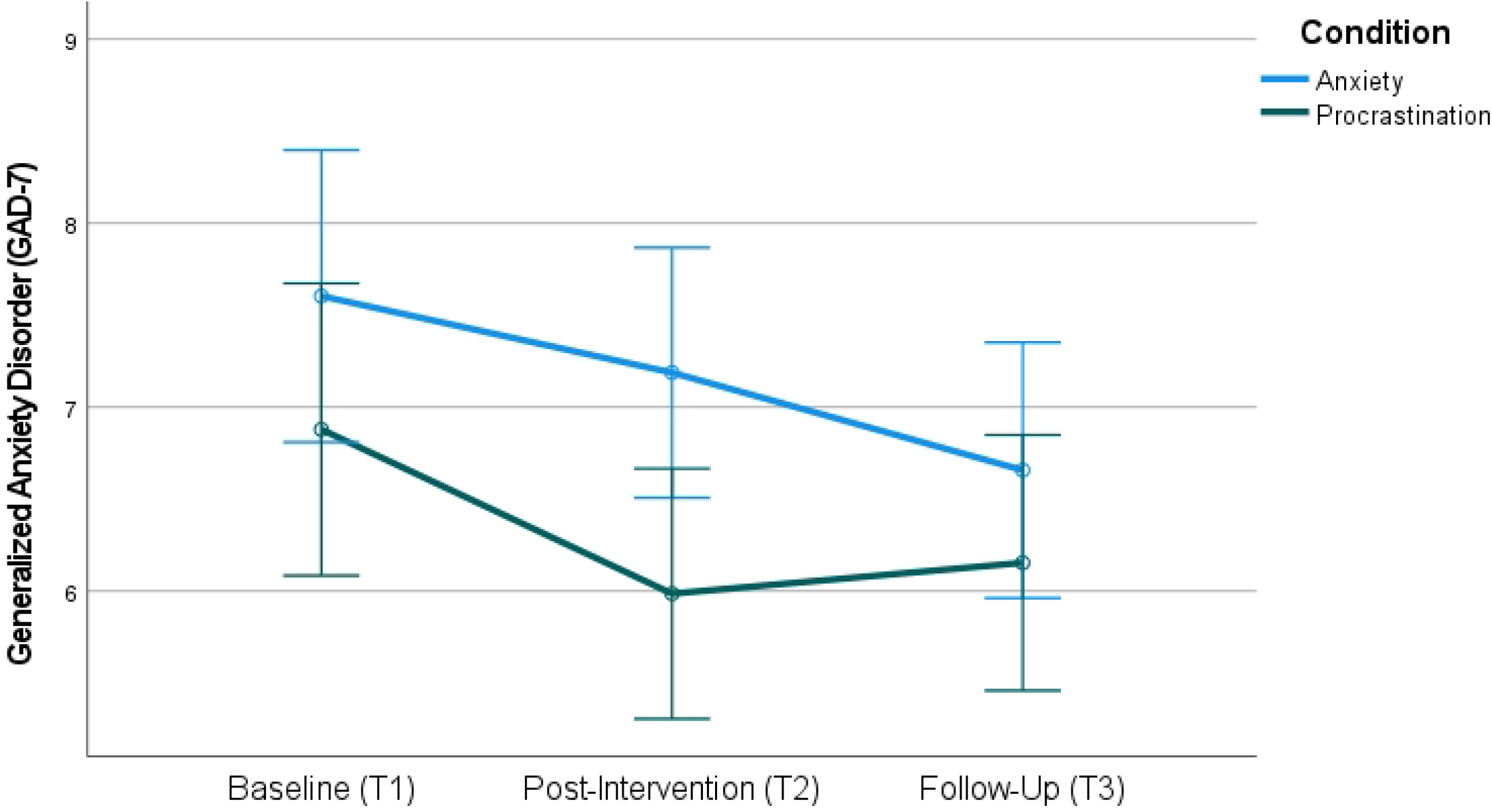
Profile Plot for changes on the GAD-7 for both groups across timepoints.

### Mediation Analyses

#### Anxiety

Hierarchical regressions controlling for condition and baseline scores found that App Engagement significantly predicted a reduction in Anxiety scores at post-intervention (ΔF (4, 196) = 43.708, p= .000, ΔR^2^= .472) and follow-up (ΔF (4, 196) = 38.798, p= .000, ΔR^2^= .442). A significant, indirect effect of PM on the relationship between App engagement and Anxiety was observed at post-intervention, β = -.049, 95% CI [-.093, -.015]. The lack of direct effects suggest that PM partially mediated the relationship between App engagement and Anxiety at post-intervention (52,53). No mediation was observed at follow-up.

#### Depression

App Engagement also significantly predicted a reduction in Depression scores at post-intervention (ΔF (4, 196) = 41.890, p= .000, ΔR^2^= .461) and follow-up (ΔF (4, 196) = 43.035, p= .000, ΔR^2^= .468). Across both timepoints, a significant mediation effect of PM was observed between App Engagement and Depression, with a standardized indirect effect of β = -.058, 95% CI [-.121, -.015] at post-intervention, and β = -.042, 95% CI [-.085, -.011] at follow-up. This means that there was a partial mediating effect of PM in the relationship between App Engagement and Depression scores in each time point. The full results are illustrated in Fig 3. A summary of the presence of direct, indirect, and total effects from all regressions is presented in Table 6.

**Table 6.**
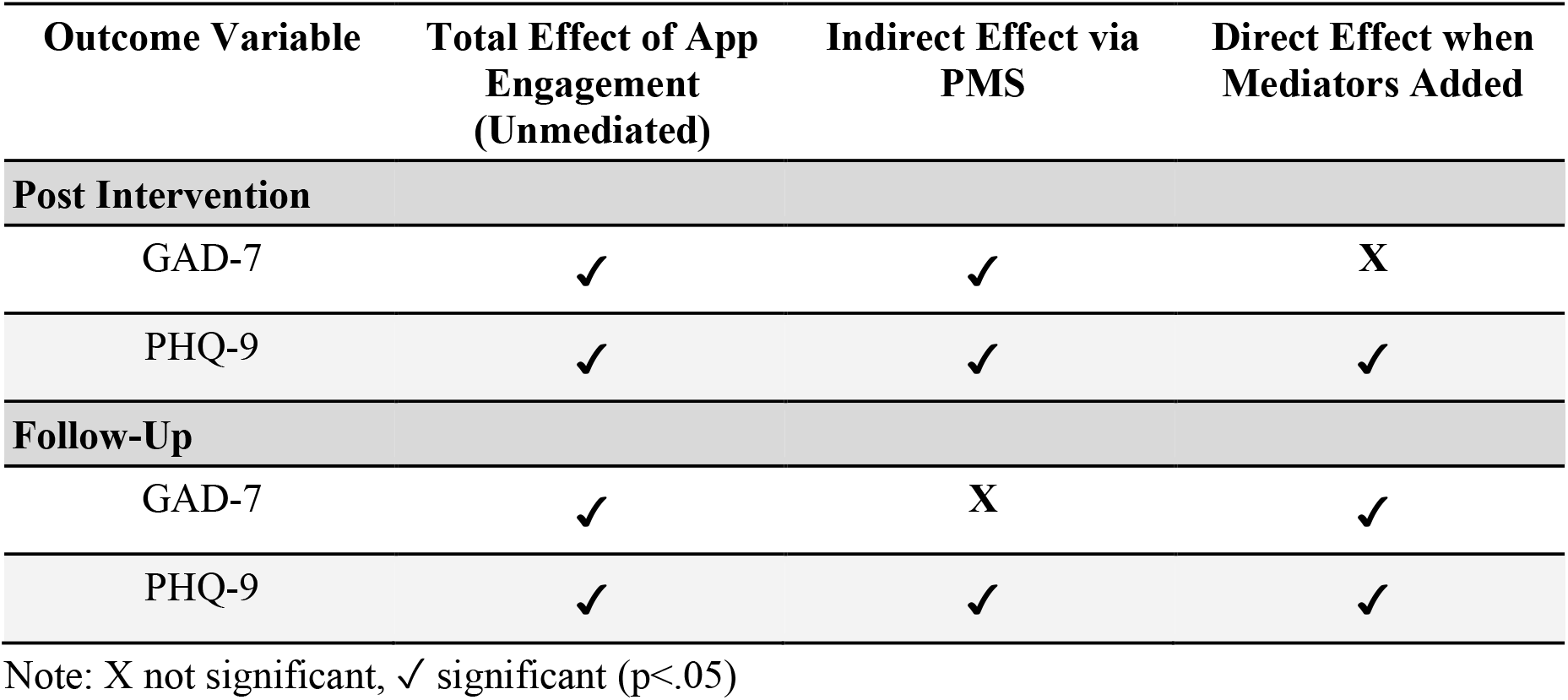
Presence of total, direct, and indirect effects of app engagement on each outcome variable.

| Outcome Variable | Total Effect of App Engagement (Unmediated) | Indirect Effect via PMS | Direct Effect when Mediators Added |
| --- | --- | --- | --- |
| **Post Intervention** |  |  |  |
| GAD-7 | ✓ | ✓ | X |
| PHQ-9 | ✓ | ✓ | ✓ |
| **Follow-Up** |  |  |  |
| GAD-7 | ✓ | X | ✓ |
| PHQ-9 | ✓ | ✓ | ✓ |

Note: X not significant, ✓ significant ( $p < .05$ )

**Fig 3.**
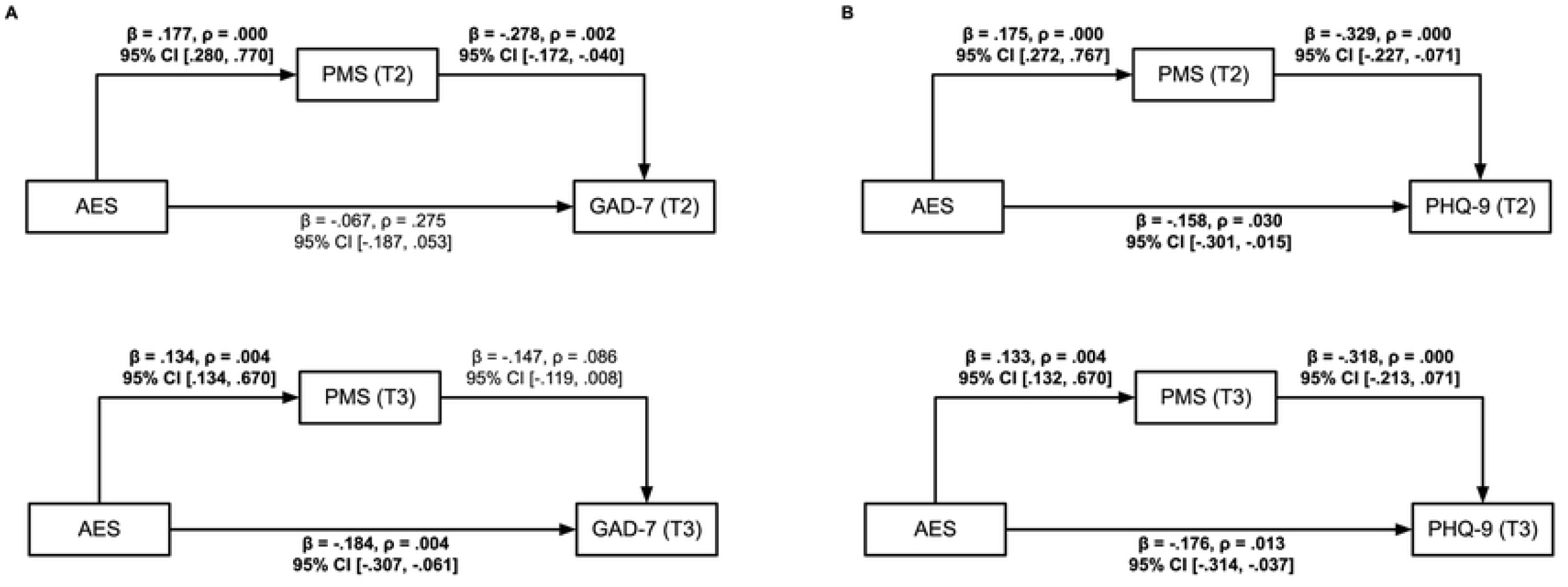
Mediated regression models using App Engagement Ratings as the predictor and (A) Anxiety and (B) Depression scores as the outcome. Whole sample (N=323) mediated regression models using App Engagement ratings as the predictor and (A) Anxiety (GAD-7), (B) Depression (PHQ-9) scores as the outcome, where T2 indicates post-intervention and T3 indicates 2-week follow-up. Note: bolded coefficients indicate significance, p<.05

## Discussion

This study sought to evaluate the effectiveness of Intellect’s mHealth programme “Anxiety and Worry” in improving anxiety and depression in college students. The tools implemented in the intervention are based on cognitive-behavior therapy principles, and include cognitive, behavioral, and writing activities that may increase an individual’s ability to self-reflect and introspect. Thus, we also examined PM as mediators of app engagement on anxiety and depression.

### Efficacy of the Anxiety and Worry Programme

As predicted, participants in the intervention group experienced a significant reduction in their anxiety levels. This is in accordance with similar studies which found that the use of mHealth applications over a period of 14 days showed a significant decrease in anxiety and depression (13,21,22). However, contrary to our hypothesis, both groups did not differ on improvements in anxiety and depression from baseline (T1) to post-intervention (T2). Interestingly, anxiety levels continued to drop further after the completion of the “Anxiety and Worry” programme, i.e. from post-intervention (T2) to follow-up (T3), whereas no such continued improvement was observed in the active control group. Similar follow-up studies also found that anxiety levels among individuals in the intervention group continued to improve after the intervention ceased (54,55). One possible explanation for this observation is Rachman’s (2016) notion that the full benefits of CBT-based intervention on anxiety will only manifest post-intervention, after participants fully understood the skills taught and practiced applying them in their day-to-day lives. Even though the intervention group was not superior to the control group in terms of lowering anxiety and depression, it is encouraging that both groups reported a significant decline in anxiety and depression scores at post-intervention and follow-up. While we cannot rule out the effect of third variables, it is plausible that both mHealth application programmes are effective in reducing anxiety and depression.

One explanation for the unexpectedly good outcomes in the active control group is that the procrastination mHealth programme is also based on the cognitive-behavior model and thus contains multiple techniques that may be beneficial for anxiety. For example, one module in the programme is titled “Overcoming the barrier” and contains discomfort intolerance related information and relaxation exercises targeting participants’ abilities to overcome difficult scenarios. The aim was to equip participants with the necessary skills to overcome similar scenarios in the future by teaching them to be mindful and walking them through how to challenge procrastination. This is supported by studies that suggest that discomfort intolerance is central to anxiety pathology (56). Studies found that discomfort avoidance, a component of discomfort intolerance, is primarily responsible for anxious responding (57). Therefore, it is likely that this module helped participants to improve discomfort intolerance and thus decreased anxiety. Moreover, this module incorporates exposure tasks related to procrastination. Peris et al. (58) previously highlighted that exposure tasks in CBT treatment were often followed by significant improvement in the rate of progress in treatment, including the primary treatment outcome (i.e. procrastination) and secondary treatment outcome (i.e. anxiety). Taken together, this suggests practical implications for future developments of mHealth apps targeting anxiety, in terms of including modules that address discomfort intolerance related to anxious responding and incorporating elements of exposure tasks.

Furthermore, helping students with procrastination may have inadvertently improved worry more than we had anticipated. Previous studies have indicated that 50-70% of college students showed frequent procrastination behavior (59) and linked procrastination to negative wellbeing outcomes, including higher anxiety and depression (60). Therefore, using PP as an active control may have exerted unexpected benefits on anxiety and depression, thereby limiting the power of the study.

Nevertheless, it is encouraging that participants in the intervention condition continued to improve post-intervention, with a steeper improvement than the one observed from baseline (T1) to post-intervention (T2), while no such benefit was observed in the control group. This suggests a potential delayed effect of the Anxiety programme, where full treatment benefits were only manifested post-intervention. One explanation for the apparent delayed effect observed is the major inclusion of writing activities (>80% of activities) within the Anxiety programme. The writing activities are framed as an interactive journal (IJ) using language that elicits change strategies, thereby mimicking motivational interviewing (MI) in writing (61). The use of MI enhances a “client-as-expert” stance, which indirectly confers a sense of autonomy and induces a belief in clients to rely on their ability to address the situation they are in. Existing MI literature has attributed this mechanism as the source of delay or the sleeper effect at post-intervention (62,63). A relevant study by Westra et al. (64) found a steeper rate of worry decline at post-intervention for MI-CBT groups. Thus, using the same principles, it is possible that the MI-based IJ components in the Anxiety programme instills a “user-as-expert” stance, which reinforce the participants’ confidence in applying the techniques learnt to address anxiety and worry, ultimately improving their anxiety levels post-intervention. This suggests practical implications for future developments of anxiety related mHealth application, in terms of integrating MI-based journaling elements which could aid in long-term effective reductions of anxiety.

### Psychological Mindedness as a Mediator

To our knowledge, this is the first study that investigated Psychological Mindedness as a mediator in the context of mHealth apps. The present study demonstrated that changes in PM mediated the relationship between higher App Engagement and improvements in anxiety and depression at post-intervention and follow-up. These findings extend previous studies by Bakker’s group (13,21,22) that have observed the mediating role of coping self-efficacy (CSE) on anxiety and depression. Previous studies have hinted upon the potential relationship between PM and CSE, particularly problem-focused or adaptive coping (38). Pang et al. (30) have also established that the insight component of PM significantly mediates the relationship between dysfunctional coping and depressive symptoms. Hence, it is unsurprising to find that PM plays a mediating role in the relationship between App Engagement and anxiety and depression outcomes.

However, only indirect effects via PM were found for Anxiety at post-intervention, but no direct effects. One explanation could be that the unmeasured presence of a conflicting mediator that ultimately cancels out the overall direct effect (52,65). This is highly probable, as the current study did not measure Coping Self-Efficacy and Emotional Self-Awareness, two previously established mediators (13,21,22). Furthermore, the present finding also supported the aforementioned theory on the potential reason behind the observed delayed effect in Anxiety as well as the importance of discomfort tolerance in anxiety pathology. A study by Beitel et al. (27) found associations between high PM individuals and their ability to tolerate distressing elements of psychotherapy. Additionally, Beitel et al. (66) also found that highly psychologically minded individuals appear to expect more self-involvement in the treatment process and outcome. Hence, speculatively, it may be that individuals who engaged with the application experienced improvements in PM, which in turn allowed them to better tolerate discomfort and become intrinsically motivated to engage with the app’s “user-as-expert”-inducing features.

A partial mediating effect of PM was found for depression at post-intervention and follow-up. This indicates that the mediating effect of PM appears to be stronger (and more apparent) in reducing depression, compared to anxiety. One reason could be the rudimentary differences between depression and anxiety; depression is typically a result of goal loss (67), resulting in persistent analytical thinking that focuses on lost goals (68). These cognitive patterns are associated with anhedonia, an inability to experience positive affect, which in turn reduces motivation to engage in other activities. On the contrary, anxiety typically surfaces as a defense mechanism when individuals are faced with threats (67). As such, a major function underlying anxiety involves selective attention to potential perceivable threats (69), whereby when faced with threatening scenarios, participants are likely to switch their attentional focus to avoid the source of threat entirely. Consequently, it is possible that PM is more relevant for depression, as its relation to motivation, rather than attentional avoidance, may have an effect in depression reduction (37). This is supported by studies that found PM to be associated with higher commitment and stronger involvement in therapy (28,70,71), which is suggestive of an individual’s motivation. Therefore, it is likely that an increase in PM influenced participants’ motivation levels, leading to incitements to produce new goals, thereby exerting a stronger mediating effect at post-intervention and follow-up. However, further studies on associations between motivation, PM and depression are required to confirm this theory.

### Limitations and Future Directions

Our study is not without limitations. First, no waitlist group was included. Although our main objective was to assess whether the Anxiety and Worry programme is more effective than the active control (Procrastination programme) in reducing anxiety and worry, including a passive control would have yielded a clearer result pattern. Furthermore, we underestimated the relevance of CBT techniques utilized in the Procrastination programme for worry outcomes and thus did not expect that it would be as effective in reducing anxiety among young adults. Alternatively, future studies may consider utilizing mHealth programmes that are not based on CBT or based on different treatment modalities. Moreover, future studies may benefit from including a more objective app engagement measure to limit its potential in acting as a third variable when examining mHealth application’s efficacy.

Lastly, although the present study provides a valuable insight into potential pathways of improvements in anxiety and depression levels via psychological mindedness when engaging in mHealth applications, future research should aim to identify additional mediators that might strengthen the benefit of mHealth applications.

## Conclusion

The intervention and active control showed significant improvements in anxiety across time, however both groups did not differ significantly from one another. Nonetheless, our study demonstrated the continual improvement of anxiety induced by the Anxiety programme post-intervention. Altogether, this study suggests that engagement with mHealth apps is related to reduction in anxiety and depression. One mechanism involved in exerting this effect is the app’s ability to improve an individual’s Psychological Mindedness. Higher engagement with either mHealth application predicted improvements on anxiety and depression scores. Despite small effect sizes observed, the practical implications of these cost-effective, highly accessible mHealth applications can meaningfully impact public mental health at scale.

## Data Availability

The data that support the findings of this study are available on request from the corresponding author (OS).

## Acknowledgements

The authors would like to thank the students for participating in the trial, the funding listed, and the team at Intellect for adapting the application and providing support throughout the study.

## Supporting Information

**S1 File. Primary and Secondary Outcomes Questionnaire**

**S2 File. Ethics Approval Letter**

